# Association between early arterial oxygenation and neurological outcome after pediatric out-of-hospital cardiac arrest: a multicenter retrospective study

**DOI:** 10.64898/2026.07.28.26359170

**Authors:** Taro Moriwaki, Shunsuke Amagasa, Masahiro Kashiura, Hideto Yasuda, Yuki Kishihara, Satoko Uematsu, Takashi Moriya

## Abstract

**Background:** Optimal arterial oxygen targets after return of spontaneous circulation (ROSC) in pediatric out-of-hospital cardiac arrest (OHCA) remain uncertain. We examined whether arterial oxygen tension on the first arterial blood gas after ROSC is associated with neurological or survival outcomes.

**Methods:** Using the Japanese Association for Acute Medicine OHCA Registry, we retrospectively studied pediatric patients (<18 years) with OHCA in whom ROSC was confirmed at or after hospital arrival. Patients were categorized by PaO₂ on the first arterial blood gas after ROSC as normoxemia (60–200 mmHg) or hyperoxemia (>200 mmHg). Missing covariate data were handled using multiple imputation, and associations were estimated using inverse probability-weighted logistic regression. Outcomes were favorable neurological status at 30 days, defined as a Pediatric Cerebral Performance Category score of 1–3, and 30-day survival. Multiple sensitivity analyses were performed, including analyses using an alternative PaO₂ threshold, restricting the timing of PaO₂ measurement, and excluding extracorporeal cardiopulmonary resuscitation cases.

**Results:** A total of 189 patients were included (95 normoxemia, 94 hyperoxemia). A favorable neurological outcome occurred in 21 of 95 (22.1%) normoxemia and 13 of 94 (13.8%) hyperoxemia patients, and 30-day survival in 40 of 95 (42.1%) and 42 of 94 (44.7%), respectively. After weighting, covariate balance was achieved with standardized mean differences below 0.1. Hyperoxemia was not significantly associated with favorable neurological outcome (adjusted odds ratio [aOR] 0.94, 95% confidence interval [CI] 0.49–1.77) or 30-day survival (aOR 1.49, 95% CI 0.88–2.54). Sensitivity analyses yielded consistent results.

**Conclusions:** Early hyperoxemia after ROSC was not significantly associated with neurological or survival outcomes in pediatric out-of-hospital cardiac arrest. These findings suggest that a single early PaO₂ measurement may be insufficient to characterize the clinical impact of oxygen exposure after resuscitation. Future research should focus on phase-specific and individualized oxygen management incorporating serial physiological assessment.

**CLINICAL PERSPECTIVE:** **What Is New?**

- This multicenter registry study evaluated the association between the first post-ROSC PaO₂ measurement and outcomes exclusively in children with out-of-hospital cardiac arrest.
- After multiple imputation and inverse probability weighting, hyperoxemia (>200 mmHg) was not significantly associated with favorable neurological outcome or 30-day survival.
- The findings were consistent across analyses using an alternative PaO₂ threshold, restricting measurement timing, and excluding extracorporeal cardiopulmonary resuscitation cases.

**What Are the Clinical Implications?**

- A single early PaO₂ value should not be interpreted as a complete measure of post-resuscitation oxygen exposure or used alone for prognostication.
- These findings do not support permissive hyperoxemia; oxygen should continue to be titrated while avoiding hypoxemia and unnecessary hyperoxemia.
- Prospective studies with serial, standardized physiological measurements are needed to define phase-specific and individualized oxygen targets.

## INTRODUCTION

Out-of-hospital cardiac arrest (OHCA) in the pediatric population is a rare event, with an incidence of approximately 0.5–20 cases per 100,000 person-years [1,2]. Despite advances in resuscitation techniques, overall survival after pediatric OHCA remains low, ranging from 6.7– 11.0 % [3,4], and only 3.7–5.1% of patients survive with favorable neurological outcomes [3,5].

One factor influencing these poor outcomes is post-resuscitation management. Appropriate oxygenation is a crucial component of this care because both insufficient and excessive oxygen delivery may worsen brain injury. Hypoxemia aggravates cerebral hypoxia, while hyperoxemia increases oxidative stress through the generation of reactive oxygen species, suggesting that maintaining normoxemia may be important [6,7].

Evidence regarding optimal post-resuscitation oxygenation in children remains limited, whereas adult evidence includes observational studies, systematic reviews, and randomized controlled trials evaluating oxygenation strategies after ROSC. In adults, observational studies and systematic reviews have generally suggested harm from hyperoxemia [8–13], but randomized controlled trials have not shown clear outcome benefit from restrictive oxygen strategies [14–16]. These adult findings cannot be directly extrapolated to children because pediatric cardiac arrest differs in physiology, arrest etiology, and patterns of care [17–19]. In pediatric observational studies, only a few have reported significant associations between PaO₂ and outcomes, whereas most have not demonstrated a clear relationship [20–27].

The inconsistent findings in pediatric studies may partly reflect heterogeneity in both patient populations and exposure assessment. Most previous pediatric studies either combined in-hospital and out-of-hospital cardiac arrest [20,22–24,26] or included in-hospital events only [21,25,27], and none has focused exclusively on out-of-hospital cardiac arrest. In studies combining both populations, PaO₂ has been assessed at widely varying intervals after ROSC, even when measurement was intended to occur early [20]. As oxygen exposure during the initial reperfusion phase may have distinct biological relevance, whether early PaO₂ is associated with outcomes in this population remains unclear. Accordingly, we conducted a multicenter retrospective cohort study to evaluate whether PaO₂ on the first arterial blood gas after ROSC was associated with neurological and survival outcomes in children following out-of-hospital cardiac arrest. We compared outcomes between normoxemia and hyperoxemia groups while adjusting for relevant confounders.

## METHODS

### Study design and setting

This study comprised a retrospective analysis of data from the Japanese Association for Acute Medicine OHCA (JAAM-OHCA) Registry, a prospective multicenter cohort of OHCA cases. The registry prospectively and consecutively enrolled all patients with OHCA who were transported to participating hospitals between 1 June 2014 and 31 December 2020. Participating institutions were emergency and critical care centers across Japan that agreed to contribute data. Prehospital information was obtained from the All-Japan Utstein Registry maintained by the Fire and Disaster Management Agency [28]. In-hospital data were recorded at each participating institution by physicians or trained staff. The JAAM-OHCA Registry Committee pooled the pre-hospital and in-hospital data [29]. The registry protocol was approved by the Institutional Review Board at each participating center, as well as by the JAAM-OHCA Registry Committee. The requirement for written informed consent was waived owing to the use of anonymized registry data. This study is reported in accordance with the STROBE guidelines [30].

### Data Availability Statement

The data that support the findings of this study are not publicly available because they are subject to data-use restrictions imposed by the JAAM-OHCA Registry. Requests for access may be directed to the corresponding author and require review and approval by the JAAM-OHCA Registry Committee. The analytic methods are described in this article and its Supplemental Material. No additional study materials were used.

### Japanese Prehospital EMS system

In the Japanese emergency medical services (EMS) system, teams of three providers deliver prehospital care, including at least one certified Emergency Medical Technician (EMT). EMTs follow the Japanese Resuscitation Guidelines, which the Japanese Resuscitation Council developed based on International Liaison Committee on Resuscitation (ILCOR) recommendations [17,19]. During cardiopulmonary resuscitation, providers use bag-valve-mask (BVM) ventilation to deliver oxygen and support ventilation. Specially trained Emergency Life Saving Technicians (ELSTs) perform advanced airway management, including endotracheal intubation in adults and supraglottic airway insertion in both adults and children, either under physician direction or according to established protocols.

### Participants

This analysis included pediatric patients younger than 18 years with OHCA in whom ROSC was confirmed at or after hospital arrival. Patients were excluded if PaO₂ data from the first arterial blood gas measurement after ROSC were unavailable or if PaO₂ was <60 mmHg to minimize potential misclassification due to venous sampling and heterogeneity related to cyanotic congenital heart disease.

### Measurements and definitions

The primary exposure was the PaO₂ on the first arterial blood gas sample obtained after ROSC. Based on previous reports [21–25, 27], a PaO₂ of 60 – 200 mmHg was classified as normoxemia and a PaO₂ greater than 200 mmHg as hyperoxemia. The primary outcome was favorable neurological status at 30 days, assessed using the Pediatric Cerebral Performance Category (PCPC) scale [31] and defined as scores of 1 (normal), 2 (mild disability), or 3 (moderate disability). The secondary outcome was survival at 30 days.

Clinical and prehospital variables were extracted from the JAAM-OHCA Registry, including age, sex, day of week, time of day, witness status, bystander CPR, initial cardiac rhythm, arrest etiology, defibrillation, adrenaline administration, intubation, extracorporeal membrane oxygenation (ECMO), presence and interventions of an accompanying physician, time from emergency call receipt to ROSC, time from ROSC to the first PaO₂ measurement, and time from patient contact by EMS to hospital arrival. Time intervals were calculated from recorded timestamps. Implausible time intervals, defined as negative values or intervals exceeding 300 minutes, were treated as missing. Cardiac arrest was defined as loss of a palpable pulse and absence of normal respiration. Out-of-hospital cardiac arrest was defined as an event occurring outside the hospital setting. Time of day was defined according to the time of the emergency call and categorized as daytime (09:00–16:59) or night-time (17:00–08:59). Etiology was classified as cardiac, respiratory, asphyxia, trauma (including traffic accidents and falls), other intrinsic causes (e.g., cerebrovascular events or malignancy), and other extrinsic causes (e.g., hanging, drowning, or poisoning).

### Statistical analysis

Baseline characteristics were summarized with categorical variables presented as numbers and percentages and continuous variables as medians and interquartile ranges. The distribution of the time from ROSC to the first PaO₂ measurement was examined descriptively. Missing covariate data were handled using multiple imputation with 20 datasets. Imputed datasets were analyzed separately and combined using Rubin’s rules to account for imputation uncertainty [32]. Baseline characteristics are presented based on the first imputed dataset, whereas standardized mean differences were averaged across all imputed datasets.

Candidate covariates were selected from clinically relevant variables available in the JAAM-OHCA Registry, guided by resuscitation guidelines [17–19], previous literature on pediatric cardiac arrest and post-resuscitation care [20–27], clinical judgment, and a directed acyclic graph (Supplementary Figure S1). Variables considered related to early post-resuscitation oxygenation, outcomes, or both were included in the analysis. Receiving hospitals were ranked according to the number of pediatric OHCA transports during the study period and classified into three volume categories, each accounting for approximately one-third of all transported cases. Propensity scores were estimated in each imputed dataset using multivariable logistic regression, incorporating age, sex, day of week, time of day, witness status, bystander CPR, initial cardiac rhythm, arrest etiology, time from patient contact by EMS to hospital arrival, defibrillation, adrenaline administration, intubation, ECMO, presence and interventions of an accompanying physician, time from emergency call receipt to ROSC, time from ROSC to the first PaO₂ measurement, and facility-volume category.

Inverse-probability weighting based on the propensity score was applied to balance baseline covariates. Covariate balance was assessed using standardized mean differences, with values <0.1 indicating adequate balance. Associations between exposure groups (hyperoxemia vs normoxemia, defined as PaO₂ >200 mmHg vs 60–200 mmHg) and outcomes were estimated using inverse probability-weighted logistic regression, yielding population-averaged effect estimates. To account for potential within-hospital correlation, robust variance estimation was clustered by receiving hospital. Crude odds ratios were estimated using univariable logistic regression with PaO₂ group as the only covariate, without weighting or adjustment for clustering. Both crude and adjusted estimates were obtained across the multiply imputed datasets and combined using Rubin’s rules. Statistical significance was inferred when the 95% confidence interval of the odds ratio did not include 1. All analyses were performed using R version 4.5.2 (R Foundation for Statistical Computing, Vienna, Austria).

### Sensitivity analysis

Sensitivity analyses were conducted to assess the robustness of the findings. First, analyses were repeated using the same analytic approach but with an alternative threshold for hyperoxemia, comparing PaO₂ >100 mmHg with PaO₂ 60–100 mmHg, consistent with thresholds used in previous pediatric studies [21,22]. Second, a complete case analysis excluding patients with missing covariate data was performed using the same propensity score weighting and outcome modeling procedures. Third, analyses were repeated after excluding the time from ROSC to the first PaO₂ measurement from the propensity score model, because this variable may act as a collider or a variable predictive of exposure but not of outcome, rather than as a baseline confounder. Fourth, analyses were restricted to patients whose first PaO₂ measurement was obtained within 60 minutes after ROSC, to reduce exposure misclassification arising from delayed sampling. Fifth, patients who received extracorporeal cardiopulmonary resuscitation (ECPR) were excluded. ECPR was defined as initiation of extracorporeal circulation before the first ROSC, whereas ECMO initiated after the first ROSC was recorded as a post-ROSC covariate. Because extracorporeal circulation directly determines arterial oxygenation through the oxygenator, the first PaO₂ value in patients undergoing ECPR may reflect circuit settings rather than endogenous oxygenation.

## RESULTS

### Patients

The linked JAAM-OHCA and All-Japan Utstein dataset included 60,349 cases. Among these, 1,241 patients were younger than 18 years, and 334 had ROSC confirmed at or after hospital arrival. A further 145 patients were excluded, including 93 with missing PaO₂ data and 52 with PaO₂ <60 mmHg, resulting in 189 patients for the final analysis. Of these, 95 were classified as normoxemic and 94 as hyperoxemic. The final analysis included 189 patients from 57 facilities, with a median of 2 patients per facility (interquartile range 2–4). The study population selection process is summarized in Figure 1. The demographics and baseline characteristics of the 93 patients excluded due to missing PaO₂ data are presented in Supplementary Table S1.

**Figure 1.**
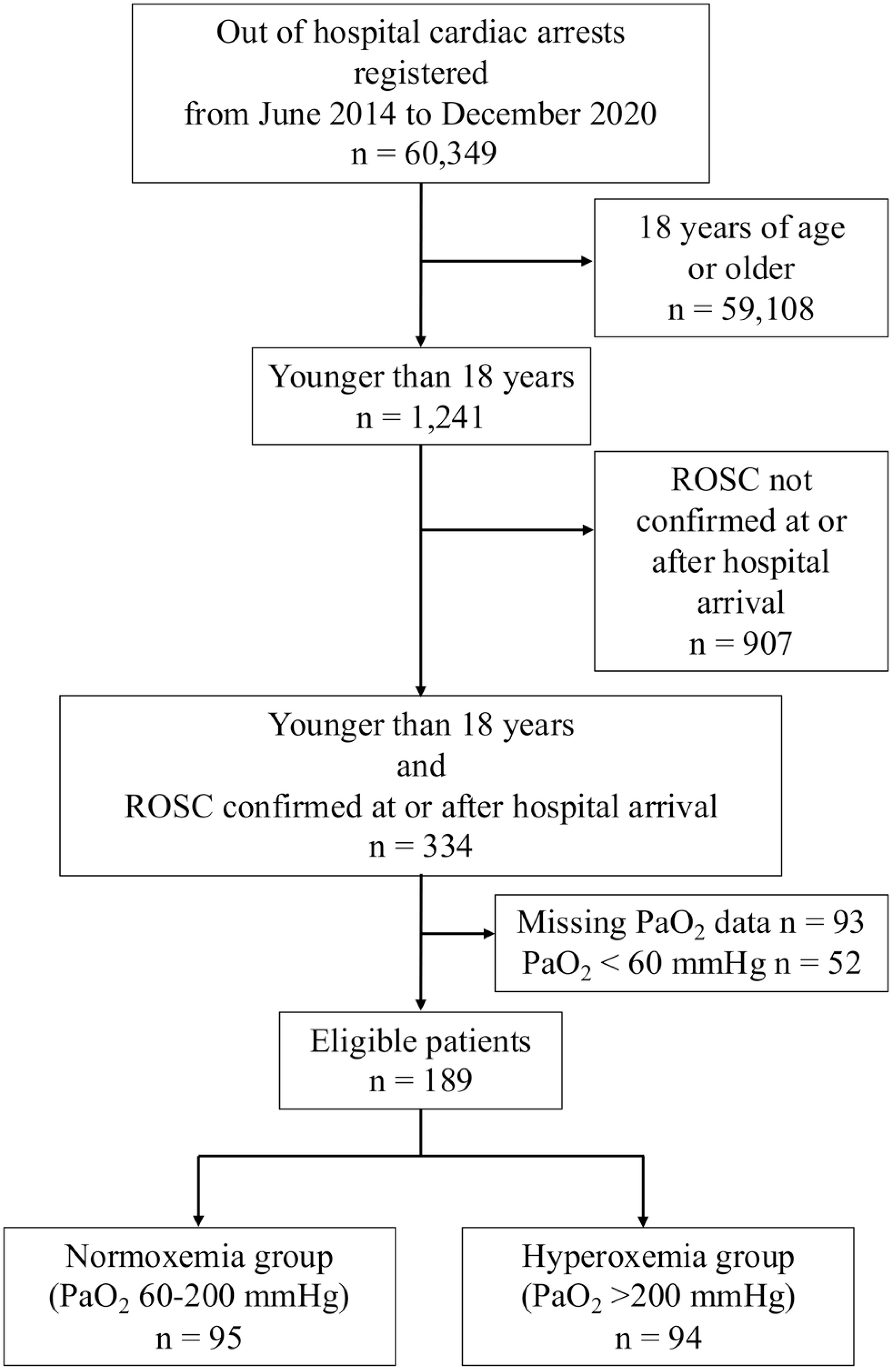
Flowchart of the study

### Demographics and patient characteristics of the study population

The overall cohort comprised 189 patients. The median age was 8.0 years (interquartile range [IQR] 1.0–15.0). Survival at 30 days was 82/189 (43.4%), whereas favorable neurological outcomes were observed in 34/189 (18.0%). Cardiac and other extrinsic causes were the most frequent etiological categories in the study population (Table 1). Among patients with available timing data (n = 182), the first arterial blood gas was obtained at a median of 24 minutes after ROSC (interquartile range 13–46 minutes), with a maximum interval of 294 minutes. As shown in Supplementary Figure S2, most measurements were obtained within 60 minutes of ROSC, and the distributions were broadly similar between the hyperoxemia and normoxemia groups.

**Table 1.** Demographic and clinical characteristics of the original cohort.

|  | Normoxemia<br>(n = 95) | Hyperoxemia<br>(n = 94) | Standardized<br>difference |
| --- | --- | --- | --- |
| Age, median (IQR) | 11 (1.0–15.0) | 7 (1.0–15.0) | 0.138 |
| Male, n (%) | 67 (70.5) | 67 (71.3) | 0.017 |
| Day of week, n (%) |  |  | 0.038 |
| Weekday | 63 (66.3) | 64 (68.1) |  |
| Weekend | 32 (33.7) | 30 (31.9) |  |
| Time of day, n (%) |  |  | 0.122 |
| Daytime (09:00–16:59) | 40 (42.1) | 34 (36.2) |  |
| Night-time (17:00–08:59) | 55 (57.9) | 60 (63.8) |  |
| Witness status, n (%) |  |  | 0.032 |
| Presence | 47 (49.5) | 45 (47.9) |  |
| Absence | 48 (50.5) | 49 (52.1) |  |
| Bystander CPR, n (%) |  |  | 0.015 |
| Presence | 63 (66.3) | 63 (67.0) |  |
| Absence | 32 (33.7) | 31 (33.0) |  |
| Initial monitored cardiac rhythm, n (%) |  |  | 0.004 |
| Shockable rhythm | 12 (12.6) | 12 (12.8) |  |
| Non-shockable rhythm | 83 (87.4) | 82 (87.2) |  |
| Cause of cardiac arrest, n (%) |  |  | 0.130 |
| Cardiac | 25 (26.3) | 24 (25.5) |  |
| Respiratory | 6 (6.3) | 5 (5.3) |  |
| Asphyxia | 13 (13.7) | 15 (16.0) |  |
| Traumatic | 11 (11.6) | 8 (8.5) |  |
| Other intrinsic | 15 (15.8) | 15 (16.0) |  |
| Other extrinsic | 25 (26.3) | 27 (28.7) |  |
| Time from patient contact by EMS to hospital arrival, median min (IQR) | 31.0 (24.0–42.0) | 30.0 (25.0–40.0) | 0.196 |
| Adrenaline administration, n (%) | 65 (68.4) | 78 (83.0) | 0.344 |
| Shock delivery, n (%) | 6 (6.3) | 13 (13.8) | 0.252 |
| Endotracheal intubation, n (%) | 84 (88.5) | 86 (91.5) | 0.115 |
| Physician during emergency transport,<br>n (%) | 13 (13.7) | 14 (14.9) | 0.035 |
| Prehospital advanced life support by<br>physician, n (%) | 24 (25.3) | 21 (22.3) | 0.069 |
| Extracorporeal membrane oxygenation<br>CPR, n (%) | 3 (3.2) | 12 (12.8) | 0.361 |
| Facility case-volume category, n (%) |  |  | 0.348 |
| Low-volume facility | 34 (35.8) | 27 (28.7) |  |
| Middle-volume facility | 36 (37.9) | 27 (28.7) |  |
| High-volume facility | 25 (26.3) | 40 (42.6) |  |
| Time from emergency call to ROSC,<br>median min (IQR) | 38.0 (27.0–<br>48.0) | 37.0 (25.0–50.0) | 0.147 |
| Time from ROSC to the first PaO<sub>2</sub><br>measurement, median min (IQR) | 23.0 (12.0–<br>46.0) | 26.0 (15.0–48.0) | 0.110 |

Values are presented as n (%) for categorical variables and median (interquartile range) for continuous variables.
IQR, interquartile range; EMS, emergency medical service; CPR, cardiopulmonary resuscitation; ROSC, return of spontaneous circulation.

After inverse probability weighting, covariate balance between exposure groups was adequate, with all standardized mean differences below 0.1 (Table 2). Crude and inverse probability-weighted outcome proportions for favorable neurological outcome and 30-day survival are presented in Table 3.

**Table 2.** Weighted baseline characteristics after inverse probability weighting.

|  | Normoxemia<br>(n = 187.8) | Hyperoxemia<br>(n = 190.6) | Standardized<br>difference |
| --- | --- | --- | --- |
| Age, median (IQR) | 10.0 (1.0–14.9) | 8.0 (1.0–15.6) | 0.013 |
| Male, % | 71.4 | 69.5 | 0.042 |
| Day of week, % |  |  | 0.046 |
| Weekday | 69.4 | 67.2 |  |
| Weekend | 30.6 | 32.8 |  |
| Time of day, % |  |  | 0.012 |
| Daytime (09:00–16:59) | 37.6 | 38.2 |  |
| Night-time (17:00–08:59) | 62.4 | 61.8 |  |
| Witness status, % |  |  | 0.042 |
| Presence | 46.6 | 48.7 |  |
| Absence | 53.4 | 51.3 |  |
| Bystander CPR, % |  |  | 0.013 |
| Presence | 69.0 | 68.4 |  |
| Absence | 31.0 | 31.6 |  |
| Initial monitored cardiac rhythm, % |  |  | 0.018 |
| Shockable rhythm | 14.1 | 14.7 |  |
| Non-shockable rhythm | 85.9 | 85.3 |  |
| Cause of cardiac arrest, % |  |  | 0.085 |
| Cardiac | 24.9 | 28.0 |  |
| Respiratory | 5.9 | 5.5 |  |
| Asphyxia | 14.2 | 14.9 |  |
| Traumatic | 9.8 | 9.2 |  |
| Other intrinsic | 15.1 | 14.7 |  |
| Other extrinsic | 30.2 | 27.6 |  |
| Time from patient contact by EMS to hospital arrival, median min (IQR) | 29.0<br>(24.0–40.0) | 30.0<br>(25.0–40.0) | 0.011 |
| Adrenaline administration, % | 75.1 | 74.4 | 0.016 |
| Shock delivery, % | 10.3 | 10.4 | 0.004 |
| Endotracheal intubation, % | 89.7 | 88.2 | 0.047 |
| Physician during emergency transport, % | 14.0 | 14.4 | 0.012 |
| Prehospital advanced life support by physician, % | 24.3 | 24.5 | 0.005 |
| Extracorporeal membrane oxygenation CPR, % | 7.4 | 7.8 | 0.016 |
| Facility case-volume category, % |  |  | 0.055 |
| Low-volume facility | 34.2 | 31.6 |  |
| Middle-volume facility | 33.0 | 34.6 |  |
| High-volume facility | 32.9 | 33.7 |  |
| Time from emergency call to ROSC, median min (IQR) | 38.0<br>(27.7–48.0) | 35.0<br>(23.5–48.5) | 0.039 |
| Time from ROSC to the first PaO<sub>2</sub> measurement, median min (IQR) | 23.0<br>(11.0–46.0) | 24.0<br>(15.0–48.0) | 0.043 |

Values are weighted percentages unless otherwise indicated; continuous variables are presented as weighted median (interquartile range).
IQR, interquartile range; EMS, emergency medical service; CPR, cardiopulmonary resuscitation; ROSC, return of spontaneous circulation.

**Table 3.** Crude and inverse probability-weighted outcome proportions.

|  | No. of patients with outcome |  |
| --- | --- | --- |
|  | Normoxemia | Hyperoxemia |
| Original cohort | n = 95 | n = 94 |
| Favorable neurological outcome at 1 month<br>after cardiac arrest, n (%) | 21 (22.1) | 13 (13.8) |
| Survival at 1 month after cardiac arrest, n (%) | 40 (42.1) | 42 (44.7) |
| Inverse probability weighting cohort | n = 187.1 | n = 190.7 |
| Favorable neurological outcome at 1 month<br>after cardiac arrest, n (%) | 36.7 (19.6) | 35.3 (18.5) |
| Survival at 1 month after cardiac arrest, n (%) | 73.5 (39.3) | 93.5 (49.0) |

### Adjusted associations between PaO₂ group and outcomes

In inverse probability-weighted logistic regression with robust variance estimation clustered by receiving hospital, compared with normoxemia, hyperoxemia was not significantly associated with favorable neurological outcome (adjusted odds ratio [aOR] 0.94, 95% confidence interval [CI] 0.49–1.77) or 30-day survival (aOR 1.49, 95% CI 0.88–2.54) (Table 4).

**Table 4.**
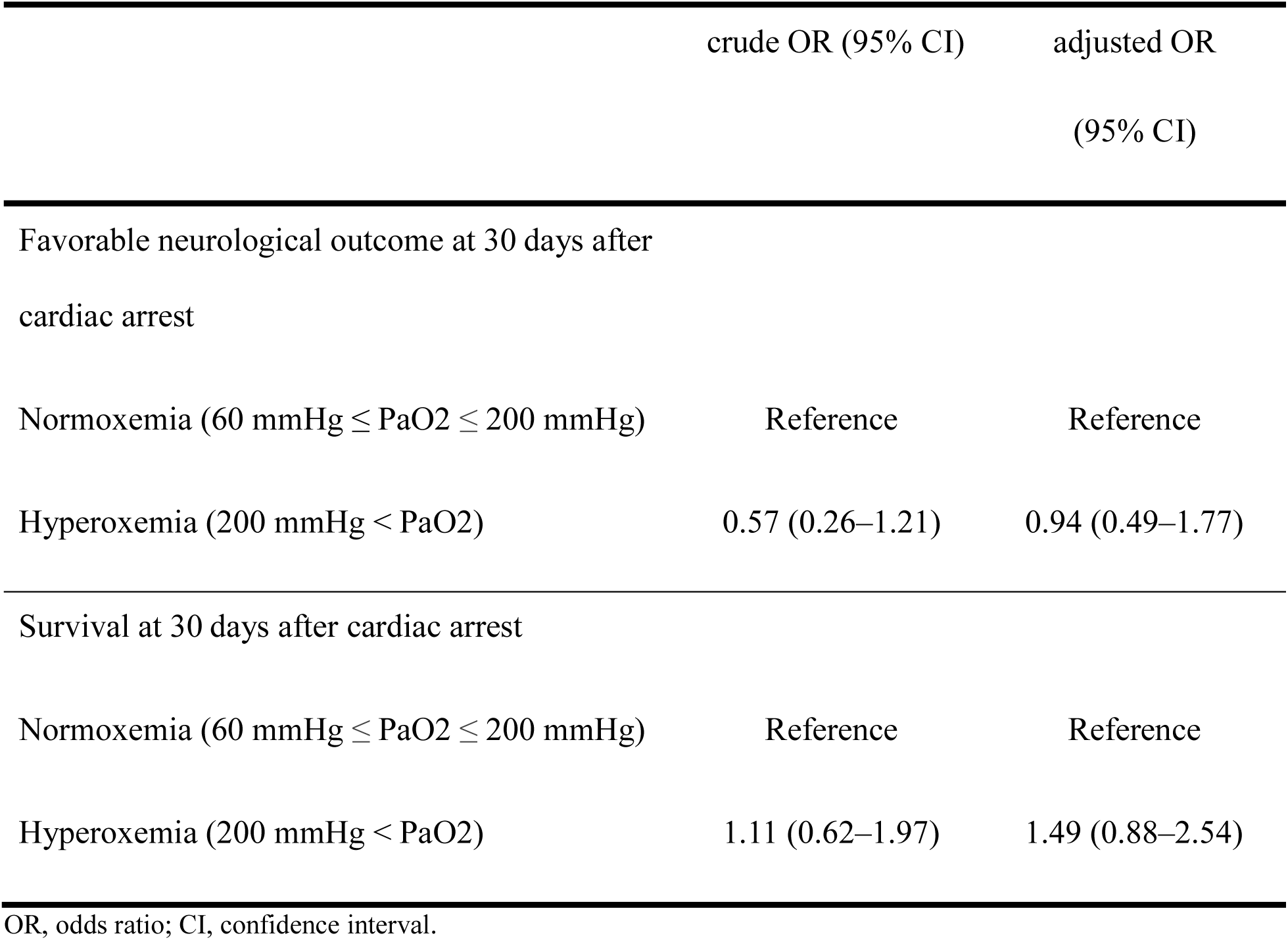
Crude and inverse probability-weighted odds ratios comparing hyperoxemia with normoxemia.

|  | crude OR (95% CI) | adjusted OR<br>(95% CI) |
| --- | --- | --- |
| Favorable neurological outcome at 30 days after<br>cardiac arrest |  |  |
| Normoxemia ($60 \text{ mmHg} \leq \text{PaO}_2 \leq 200 \text{ mmHg}$) | Reference | Reference |
| Hyperoxemia ($200 \text{ mmHg} < \text{PaO}_2$) | 0.57 (0.26–1.21) | 0.94 (0.49–1.77) |
| Survival at 30 days after cardiac arrest |  |  |
| Normoxemia ($60 \text{ mmHg} \leq \text{PaO}_2 \leq 200 \text{ mmHg}$) | Reference | Reference |
| Hyperoxemia ($200 \text{ mmHg} < \text{PaO}_2$) | 1.11 (0.62–1.97) | 1.49 (0.88–2.54) |

OR, odds ratio; CI, confidence interval.

### Sensitivity analyses

Sensitivity analyses using an alternative definition of hyperoxemia (PaO₂ >100 mmHg) yielded similar adjusted associations for both outcomes (Table 5, Supplementary Tables S2–S4). Complete-case analyses excluding patients with missing covariate data also showed results consistent with the primary analysis (Supplementary Tables S5–S8). In addition, the results were unchanged when the time from ROSC to the first PaO₂ measurement was excluded from the propensity score model (Supplementary Tables S9–S12). Restricting the analysis to patients whose first PaO₂ was measured within 60 minutes of ROSC yielded consistent results (Supplementary Tables S13–S16). Excluding patients who received ECPR also did not materially change the associations (Supplementary Tables S17–S20).

**Table 5.** Crude and inverse probability-weighted odds ratios comparing hyperoxemia with normoxemia in the sensitivity analysis using a PaO₂ threshold of 100 mmHg.

|  | crude OR (95% CI) | adjusted OR<br>(95% CI) |
| --- | --- | --- |
| Favorable neurological outcome at 30 days after cardiac arrest |  |  |
| Normoxemia (60 mmHg ≤ PaO<sub>2</sub> ≤ 100 mmHg) | Reference | Reference |
| Hyperoxemia (100 mmHg < PaO<sub>2</sub>) | 0.57 (0.26–1.27) | 0.52 (0.21–1.26) |
| Survival at 30 days after cardiac arrest |  |  |
| Normoxemia (60 mmHg ≤ PaO<sub>2</sub> ≤ 100 mmHg) | Reference | Reference |
| Hyperoxemia (100 mmHg < PaO<sub>2</sub>) | 1.15 (0.60–2.23) | 1.28 (0.65–2.54) |

OR, odds ratio; CI, confidence interval.

## DISCUSSION

In this multicenter pediatric out-of-hospital cardiac arrest cohort in Japan, early hyperoxemia, defined as a PaO₂ greater than 200 mmHg on the first arterial blood gas obtained after return of spontaneous circulation, was not significantly associated with favorable neurological outcomes or 30-day survival after propensity score weighting for measured prehospital and in-hospital covariates (adjusted odds ratio 0.94, 95% CI 0.49–1.77 for favorable neurological outcome; 1.49, 95% CI 0.88–2.54 for survival). Sensitivity analyses using an alternative PaO₂ threshold, complete-case analyses, models excluding the time from ROSC to the first PaO₂ measurement from the propensity score, analyses restricted to patients whose first PaO₂ was measured within 60 minutes of ROSC, and analyses excluding patients who received ECPR all yielded consistent results.

The association between post-resuscitation PaO₂ and outcomes has been inconsistently reported across pediatric studies [20–27]. One large multicenter cohort of children admitted to pediatric intensive care units reported higher mortality among those with severe hyperoxemia [20]. However, that study included both out-of-hospital and in-hospital cardiac arrests, assessed PaO₂ after ICU admission rather than early after ROSC, and used mortality rather than neurological outcome as the primary endpoint. These design differences limit direct comparability with the present findings. Most other pediatric studies that assessed PaO₂ closer to the time of ROSC have not demonstrated a significant association with neurological or survival outcomes [21–27], although the timing of measurement varied substantially across studies, such that “early” PaO₂ values likely reflected heterogeneous physiological phases of post-arrest care. Taken together, the existing pediatric literature has not demonstrated a consistent association between early PaO₂ and clinical outcomes.

A key source of heterogeneity across prior studies is the variation in how oxygen exposure has been operationalized. While some studies assessed single PaO₂ values at variable time points after ROSC [20–23, 25, 27], others used time-weighted metrics reflecting cumulative exposure [24,26]. Cumulative metrics may more comprehensively reflect the dynamic trajectory of oxygenation, but they require frequent and standardized sampling, the availability of which varies across patients and institutions. They may also be susceptible to informative sampling and survival-related biases. In the present study, we focused on the first PaO₂ measurement after ROSC as a consistent and clinically interpretable indicator of early oxygen status. This approach prioritizes clinical reproducibility and reflects a clinically available early measurement, although it cannot account for the duration of hyperoxemic exposure or subsequent changes in oxygenation.

During the initial reperfusion phase after ROSC, cerebral tissue may be particularly vulnerable to hyperoxemia because of rapid reactive oxygen species generation, depletion of antioxidant reserves, and hyperoxia-induced cerebral vasoconstriction that may reduce cerebral oxygen delivery [33–36]. In adult studies, hyperoxemia within the first hours after ROSC has been associated with worse neurological and survival outcomes [10,13]. In the present study, the first PaO₂ was obtained at a median of 24 minutes after ROSC, with most measurements obtained within 60 minutes, representing one of the earliest assessments in the pediatric literature. Nevertheless, no significant association with either outcome was identified. In the analysis restricted to measurements within 60 minutes of ROSC, the adjusted odds ratio for survival was somewhat higher (1.76, 95% CI 0.96–3.21) but remained non-significant, likely reflecting the smaller subgroup size.

The first PaO₂ measurement after ROSC may also have influenced subsequent oxygen titration and ventilatory management. Recognition of hyperoxemia may have prompted clinicians to reduce inspired oxygen concentrations, thereby attenuating cumulative oxygen exposure in the hyperoxemia group, whereas later hyperoxemic episodes in initially normoxemic patients may have been missed. Such downstream treatment modification may have reduced separation in oxygen exposure between groups and attenuated potential associations with outcomes.

These findings highlight important considerations for future research on post-resuscitation oxygen management in pediatric out-of-hospital cardiac arrest. Although the early reperfusion phase after ROSC is biologically important, it likely represents a heterogeneous physiological period rather than a uniform exposure window. PaO₂ measured within minutes after ROSC may reflect different pathophysiology from values obtained several hours later, when shock progression, respiratory dysfunction, and therapeutic interventions may already have altered oxygen delivery and utilization. Future prospective studies should therefore incorporate serial and standardized blood gas assessments during the early post-arrest period to evaluate oxygen exposure trajectories more consistently. Because oxygenation and ventilation strategies may be modified in response to measured PaO₂ values, future analyses should also account for treatment adaptation and interfacility practice variation. Achieving these aims will require sufficiently large multicenter cohorts with protocolized physiological monitoring.

However, several limitations should be acknowledged. First, despite adjustment for multiple prehospital and in-hospital variables, residual confounding cannot be excluded because detailed data on baseline comorbidities, underlying cardiac conditions, and oxygenation or ventilation strategies before the first arterial blood gas measurement were unavailable. These unmeasured factors may have influenced the initial PaO₂ value and may also have been related to clinical severity, complicating interpretation of the independent association between early PaO₂ and outcomes. Second, patients without PaO₂ measurements were excluded, which may have introduced selection bias. Excluded patients tended to be younger than those included in the primary analysis, possibly reflecting technical challenges in obtaining arterial blood gas samples in smaller children, and this may limit generalisability to younger pediatric populations.

Furthermore, exclusion of patients with PaO₂ < 60 mmHg to minimize potential misclassification due to venous sampling or cyanotic congenital heart disease may have further restricted the study population. Third, only the first arterial blood gas after ROSC was analyzed. Although most measurements were obtained within 60 minutes of ROSC (Supplementary Figure S2), variability in measurement timing and the inability to assess subsequent PaO₂ changes or cumulative oxygen exposure may have introduced exposure heterogeneity. Finally, the modest sample size limited statistical precision and reduced the ability to detect small but clinically meaningful associations.

## CONCLUSIONS

In this multicenter pediatric out-of-hospital cardiac arrest cohort in Japan, early post-resuscitation hyperoxemia was not significantly associated with favorable neurological outcome or 30-day survival. These findings suggest that a single early PaO₂ measurement, when considered in isolation, may be insufficient to characterize the clinical impact of oxygen exposure after resuscitation. Further research is needed to better define optimal oxygenation strategies after return of spontaneous circulation, including approaches that incorporate serial oxygenation assessment and dynamic physiological monitoring across the early post-arrest period while accounting for patient heterogeneity.

## ACKNOWLEDGMENTS

We gratefully acknowledge the cooperation of the members and institutions of the JAAM-OHCA Registry, listed at http://www.jaamohca-web.com/list/.

## AUTHOR CONTRIBUTIONS

Taro Moriwaki takes responsibility for the manuscript as a whole. Taro Moriwaki, Shunsuke Amagasa, Masahiro Kashiura, Hideto Yasuda, Yuki Kishihara, Satoko Uematsu, and Takashi Moriya designed the study. Taro Moriwaki and Shunsuke Amagasa analyzed the data. Taro Moriwaki drafted the manuscript. Taro Moriwaki, Shunsuke Amagasa, Masahiro Kashiura, Hideto Yasuda, Yuki Kishihara, Satoko Uematsu, and Takashi Moriya critically revised the manuscript. All authors read and approved the final version of the manuscript.

## USE OF GENERATIVE ARTIFICIAL INTELLIGENCE

During the preparation of this work the authors used ChatGPT (OpenAI) and Claude (Anthropic) in order to improve the readability and language of the manuscript. After using these tools, the authors reviewed and edited the content as needed and take full responsibility for the content of the published article.

## NONSTANDARD ABBREVIATIONS AND ACRONYMS

OHCA: out-of-hospital cardiac arrest
ROSC: return of spontaneous circulation
PaO₂: partial pressure of arterial oxygen
JAAM-OHCA: Japanese Association for Acute Medicine Out-of-Hospital Cardiac Arrest
ILCOR: International Liaison Committee on Resuscitation
EMS: emergency medical services
EMT: emergency medical technician
BVM: bag-valve-mask
PCPC: Pediatric Cerebral Performance Category
CPR: cardiopulmonary resuscitation
ECMO: extracorporeal membrane oxygenation
ECPR: extracorporeal cardiopulmonary resuscitation
IQR: interquartile range
OR: odds ratio
CI: confidence interval

